# Variation in Nutritional Composition of Anatomical Parts and Taxonomic Classes of Wild Animals: A Systematic Review Using Data Imputation with Artificial Intelligence

**DOI:** 10.1101/2024.10.22.24315931

**Authors:** Ana Luisa dos Santos Medeiros, Amanda Letícia Bezerra de Oliveira, Maria Fernanda Araújo de Medeiros, Daniel Tregidgo, Eliana Bistriche Giuntini, Elias Jacob de Menezes Neto, Juliana Kelly da Silva Maia, Michelle Cristine Medeiros Jacob

## Abstract

Wildmeat is crucial for the food security of Indigenous Peoples and Local Communities, yet information about its nutritional profile remains scarce. This study systematically reviewed the impact of anatomical part and taxonomic class (mammals, birds, reptiles) on the nutritional composition of wildmeat. Using the PRISMA protocol, we selected articles from Web of Science, Scopus, and Medline/PubMed databases, with criteria including original articles on wild animal meat composition consumed by humans, excluding studies presenting secondary data or lacking detailed methodologies. We employed a quality questionnaire and concordance analysis (Fleiss’ Kappa = 1.00) for robustness. Artificial intelligence techniques (eg., K-Nearest Neighbors) estimated missing nutritional values in all 21 articles included in our study, covering 26 species and 10 nutrients. Results show statistically significant nutritional variations between anatomical parts and animal classes. Reptile viscera have over twice the fat content and triple the iron compared to muscles. Mammal viscera contain five times more omega-6 and selenium, four times more iron and manganese, and almost double the zinc compared to muscles. Among classes, bird muscles have over 90% higher fat content than mammal muscles and 20% higher than reptile muscles. Mammals have over 100% higher zinc levels than birds, and reptiles have over 400% more selenium than birds. No significant difference in iron content between mammals and birds was noted, likely due to bird slaughter methods. This study highlights the importance of wildmeat for food security. Importantly, we demonstrate an enormous variation in nutritional composition, underscoring how different anatomical parts and taxonomic classes can contribute to tackling different nutritional deficiencies. Additionally, it introduces a novel methodology for handling missing nutritional composition data, providing a comprehensive approach to understanding the nutritional value of wildmeat. Our findings can inform food security policies and wildlife management, balancing conservation and subsistence.

## 1. INTRODUCTION

Meat, as a fundamental component of human diets, stands out for its diversity and nutritional value, offering an abundant source of essential nutrients for health. In this context, meat from wild animals, known as wildmeat, bushmeat or game meat, emerges as a fundamental resource for various populations around the world, especially Indigenous Peoples and Local Communities (IPLC). In this study, we consider wildmeat to be derived from wild, non-domesticated animals, (1) particularly vertebrates, excluding insects, crustaceans, larvae, mollusks, and fish. Wildmeat is a crucial resource for ensuring food access for populations within traditional food systems (2–4). It is estimated that approximately 230 to 833 million people rely on wild animal meat as a source of protein worldwide (5). Furthermore, its consumption is associated with improved nutritional status and cognitive development (6). For instance, an investigation in the Amazon region found a correlation between wildmeat consumption and children’s health, showing an average increase in hemoglobin concentration of 0.25 g/dL in children from vulnerable rural areas (7). Similarly, a study in Madagascar revealed that families most reliant on wildmeat, who are also the most economically disadvantaged, have a four-fold higher likelihood of developing anemia if they lose access to wildlife (8) .

Despite progress in recognizing the importance of wildmeat for food security and health, significant gaps remain in understanding its nutritional composition, particularly regarding micronutrients. Few studies have specifically evaluated the nutritional composition of wild species consumed (9), and existing nutritional composition tables, such as the Brazilian Food Composition Table, (10) often do not distinguish between wild and domesticated species. Researchers frequently use food matching to estimate the nutritional composition of foods with unavailable direct data, (11) a method critical for assessing products derived from biodiversity. However, this approach may introduce inaccuracies if the data is based on domestic animals, potentially misrepresenting wild species’ nutritional profiles (12).

Current literature provides limited and scattered data on the nutritional composition of wild animals, (9,13,14) hindering a comprehensive understanding of nutritional variations. Therefore, our systematic review aims to identify patterns in the nutritional composition of wildmeat, compiling data and applying artificial intelligence techniques to address missing values (15). This approach offers a detailed analysis of wild animal meat’s nutritional profile, addressing ethical challenges in studying wild species.

Currently, the scientific literature presents, albeit limited, data on the nutritional composition of wild animals (9,13,14). However, this information is scattered across various articles and diverse fields of knowledge. This dispersion and scarcity of data significantly hinder our comprehensive understanding of nutritional variations in wild animals. In light of this, we propose the systematic review we conducted to identify patterns in the nutritional composition of wildmeat. By compiling this data and applying artificial intelligence techniques to address missing values, (15) we provide a detailed analysis of the nutritional profile of wildmeat. This approach represents a viable strategy in the face of the ethical challenges involved in analyzing wild species, considering that our database contained many gaps due to the scarcity of data in the literature, likely caused by legal and ethical limitations in sample collection (16).

Different tissues, such as viscera and muscles, have distinct nutritional profiles due to their specific physiological functions (17). Additionally, animal class may determine nutritional composition variations influenced by physiological and environmental factors (2,9). In this sense, to identify nutritional patterns in wildmeat composition, we hypothesized that the anatomical part (viscera vs. muscles) and the animal class (mammals, birds, reptiles) influence meat’s nutritional composition.

## 2. MATERIALS AND METHODS

The data used to create the *corpus* for this project were gathered through a systematic review conducted according to the Preferred Reporting Items for Systematic Reviews and Meta-Analyses (PRISMA) guidelines, [(18)] see Supplementary Material 1. The protocol for this review was not registered in advance, as the research does not directly analyze any health-related outcomes.

### 2.1 Selection criteria and search

To address the research question posed in this project, “How does the nutritional composition of anatomical parts and taxonomic classes of wild animals vary?”, we selected articles based on the following eligibility criteria: (i) original articles published in any language, with no date restrictions, that (ii) provided data on the composition of wild animal meat consumed by human populations. We also included articles recommended by experts in the field, provided they met the eligibility criteria. We excluded studies that (i) did not provide nutritional composition data of wild animals, (ii) presented secondary data, or (iii) did not detail the methodology for food composition analysis.

In June 2022, we conducted searches in the Web of Science, Scopus, and Medline/PubMed databases (via the National Library of Medicine). The search involved applying specific descriptors in each database. The following strategy guided the search: (BUSHMEAT OR “WILD MEAT” OR “GAME MEAT” OR “INDIGENOUS MEAT” OR “WILD ANIMALS” OR “HUNTING ANIMALS MEAT”) AND (“FOOD CONSUMPTION” OR “FOOD INTAKE” OR DIETARY) AND (MICRONUTRIENT OR NUTRIENT OR NUTRITION OR “FOOD COMPOSITION” OR MINERAL).

### 2.2 Study selection

We used the Rayyan tool to organize records and remove duplicates identified during the search. Three authors (ALSM, MFAM, ALBO) independently screened the articles, applying the eligibility criteria. During the initial screening, titles and abstracts that did not meet the inclusion criteria were excluded. In the next stage, full-text reviews were conducted, and studies presenting secondary data, incomplete works, or not meeting the selection criteria were excluded.

In cases of discrepancies or uncertainties, two authors (MCMJ, JKSM) were consulted. Subsequently, potentially eligible articles were read in full. After the reading and selection process, the chosen articles were stored in the Zotero reference management software.

### 2.3 Data extraction

Three authors (ALSM, MFAM, ALBO) independently extracted data from the selected articles to ensure transcription accuracy. Initially, we organized article identification data and information on the method of food composition analysis used in each study to determine the comparability of the available nutritional information. Additionally, we extracted data on research variables of interest, namely: taxonomy, animal class, analyzed part, and levels of macronutrients and micronutrients.

The nutrients analyzed, given the scope of the study (e.g., excluding heavy metal analysis) and data availability, include the following: iron, selenium, zinc, potassium, magnesium, manganese, sodium, proteins, fat, and omega-6. Nutritional composition data were converted into grams per 100 grams, and all levels were standardized on a wet weight basis. Articles presenting data on a dry weight basis were individually converted and calculated using moisture information described in the methodology of the respective articles. It is noteworthy to mention that we did not have access to the raw data from the original analyses conducted by the authors of the papers reviewed, as this information was not provided.

### 2.4 Quality analysis

To date, there are no specific quality questionnaires for research on the nutritional composition of wild animals in the scientific literature. Creating such a tool is essential, considering the ethical and legal complexities associated with studying wildmeat (19). These complexities include the use of opportunistic sampling and the need for rigorous control over potential conflicts of interest in the studies analyzed. To address this gap, we developed a quality questionnaire aimed at assessing the methodological robustness of these studies.

The assessment tool consisted of a checklist with nine items. The development of this instrument followed an integrative approach, incorporating elements from existing protocols such as LatinFoods/FAO, “QUADAS Tool/Timmer’s Analysis Tool,” OHAT, Cochrane, QUADAS, Timmers, and STROBE, and adapting them to meet the specific needs of our research. For each item, we crafted a corresponding question that a well-reported study should address, covering aspects such as objectives, sample size, study design, methods, results, statistical analysis, funding sources, conflicts of interest, and incomplete data. Structured responses and scoring were developed for each question: 0 points indicate that information is not available, 0.5 points denote ambiguity or unclear information, and 1.0 point signifies that the information is clearly presented or not applicable (see Supplementary Material 2).

The tool was tested by three trained evaluators (ALSM, MFAM, and ALBO), who used the instrument to independently assess the quality of an article. Subsequently, the results from each evaluator were discussed with the research team to refine the clarity of the instrument. With the tool finalized, three trained evaluators independently assessed the articles using the checklist. The checklist data were computed individually, and an average score was calculated from the evaluators’ assessments. The agreement between the results was measured using Fleiss’ Kappa. Articles were classified as low quality if they received between 0 to 2.9 points, medium quality if they received between 3.0 to 5.9 points, and high quality if they received more than

### 6.0 points

### 2.5 Data preparation

Table 1 lists the amount of missing data for each nutrient column, considering the total of 76 observations in the dataset.

**Table 1.**
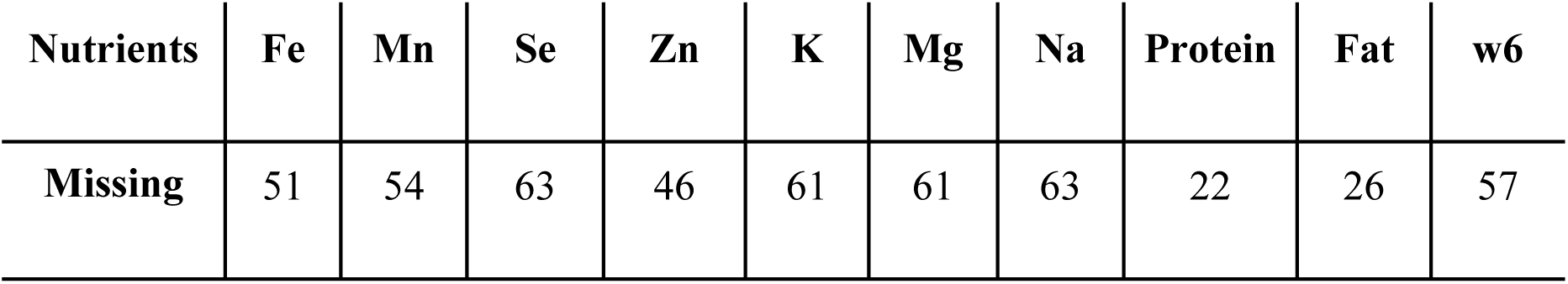
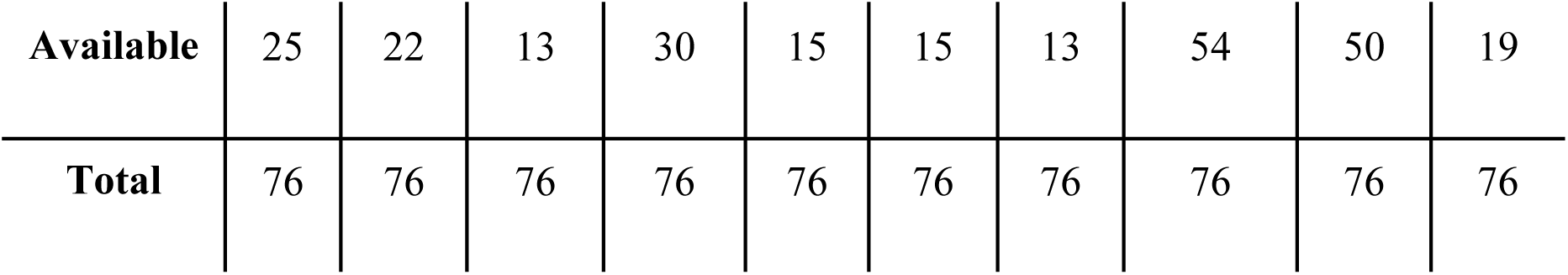
Overview of available and missing data for each nutrient analyzed.

To address these gaps, we used machine learning to impute missing data. The goal is to frame each missing value as a regression problem, estimating missing values to minimize their impact on analysis results. This method expands the use of the original data, by allowing its use on modeling steps that would not be possible with the missing value (20,21).

Generally, there are three main types of missing data: Missing Completely at Random (MCAR), Missing at Random (MAR), and Missing Not at Random (MNAR). MCAR denotes cases where the probability of a value being missing does not depend on the data itself or any other observed variables. In contrast, MAR means that the probability of missingness is related to other observed attributes. Finally, MNAR arises when the probability of a value being missing is explicitly dependent on the missing value itself (22).

In the context of our dataset, we consider the absence of values in the nutritional data as Missing Completely at Random (MCAR), indicating that the missing data are unrelated to the dataset itself. This assumption seems reasonable, as our literature review revealed that the scarcity of data is primarily attributed to the limited availability of studies on game meat analysis. This is likely justified by the legal and ethical limitations in collecting samples (16), rather than any specific bias or underlying structural cause. The missing values are likely a result of general research limitations such as funding constraints or legal and ethical constraints on sample collection. We tested several methods to impute our data, including:

1. *Multiple Imputation by Chained Equations* (MICE): This method operates iteratively, imputing missing values with estimates derived from regression models using observed data.
2. *K-Nearest Neighbors* (KNN): This technique estimates missing values based on the average of the k nearest neighbors, considering the distance between variables in the dataset.
3. *Soft Impute:* In this approach, missing values are estimated through a low-rank matrix approximation using a soft thresholding algorithm.
4. *Matrix Factorization:* This technique decomposes the matrix into low-dimensional components, which are then used to approximate missing values within the matrix.
5. *Nuclear Norm Minimization:* This method involves minimizing the nuclear norm of the matrix, resulting in a low-rank approximation of the original matrix with complete data.
6. *Iterative Singular Value Decomposition* (SVD): In this approach, an iterative algorithm is employed to estimate missing values, progressively refining the approximation using SVD.

We evaluated the best technique among these mentioned using a cross-validation approach for data imputation methods. This approach involves intentionally removing known values from the dataset, treating them as missing, and then attempting to impute them using various techniques. It is important to note that our imputation process considered the entire dataset, not just individual columns. For example, when imputing missing values in the iron (Fe) column, the process took into account all available data across all nutrient columns. This multivariate approach allows the imputation methods to leverage relationships and patterns between different nutrients, potentially improving the accuracy of the estimates. This way we can capture complex interactions between nutrients that might be missed if we only looked at each nutrient in isolation. This process allows us to compare the performance of each imputation method by estimating the reliability of the imputed values against the actual known values. The steps to execute this evaluation method are as follows:

1. Selecting a known value from the dataset and masking it, treating it as a missing value.
2. Apply each imputation technique separately to estimate the masked value.
3. Store the imputed values obtained from each method for further analyses.
4. Repeat steps 1-3 for all available values in all columns of the dataset.
5. Once the imputation process is complete for all values and methods, calculate the chosen evaluation metrics by comparing the original values with those imputed using each of the different imputation methods.

For the final step, the assessment of imputation, we used three main metrics: Symmetric Mean Absolute Percentage Error (SMAPE)and Mean Absolute Error (MAE). MAE is useful for assessing the overall performance of the imputation technique, providing a measure of how closely imputed values approximate the actual values. SMAPE quantifies the relative accuracy between observed and imputed values by calculating the average of absolute differences normalized by the average absolute values, subsequently expressed as a percentage. The formula for SMAPE is as follows:

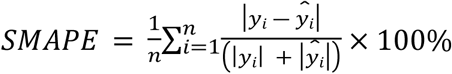

Where:

- *y*_*i*_ is the actual value of the variable
- *ŷ_i_* is the predicted value of the variable
- *n*is the number of observations

Finally, the MAE (Mean Absolute Error) calculates the average of the absolute differences between observed and imputed values, providing a direct and interpretable representation of the average error magnitude. The formula for MAE is given by:

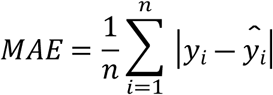

In summary, our methodology includes a thorough analysis of missing data characteristics. We classified the pattern of absence and employed a systematic execution of multiple imputation techniques to address the limitation caused by missing values. The scarcity of nutritional information for wildmeat, identified in our literature review, presents a compelling case to treat missing nutrition data as MCAR. This assumption allows for the use of imputation methods, which, as shown in previous research, can enhance the accuracy and reliability of estimates derived from incomplete datasets.

All data imputation analyses were conducted using the Python programming language. Upon completion of the imputation process, nutrient values were expressed in units of measure as per the Dietary Reference Intakes. [(23)] The code required to replicate our results is publicly available on GitHub. [(24)]

### 2.6 Variables related to hypothesis testing

The nutritional composition data of 10 nutrients served as our dependent variable. For hypothesis 1, where we tested the influence of the anatomical part on nutritional composition, the independent variables were muscle and viscera. For hypothesis 2, the independent variables were the animal classes: mammals, birds, and reptiles.

We recognize that factors such as animal anatomy (25) cause of death, (13) geographical location and seasonality, (26,27) physical activity level, (28) and physiological and metabolic characteristics (9) can affect meat’s nutritional composition. However, we lack access to these data for our research because they are inconsistently reported in the analyzed studies and are only occasionally mentioned when investigating these specific influences.

### 2.7 Data Analysis

We used descriptive statistics to summarize our findings and assessed the normality of numerical data using the Shapiro test. Based on the data characteristics, we conducted either ANOVA or Kruskal-Wallis tests, setting a significance level of 0.05 (p-value) to determine the significance of the results. In cases of significant differences, we performed post hoc tests such as Tukey or Bonferroni. All statistical analyses were conducted using the R programming language through the RStudio interface.

## 3. RESULTS

### 3.1 Study Selection

After searching the database, we obtained a total of 565 articles (Web of Science: 68, Medline/PubMed: 325, Scopus: 172). After excluding 142 duplicates, 423 articles proceeded to title and abstract screening. Following this step, we selected 75 articles for full-text review. At this stage, 62 articles were excluded for presenting secondary data (5 articles), being incomplete (56 articles), or lacking nutrient composition data relevant to our selection (1 article). Ultimately, we included 13 articles for review. Additionally, we incorporated 8 external articles, recommended by experts, which met all our criteria. Therefore, a total of 21 articles constitute our study. Figure 1 illustrates the selection process and flowchart.

**Figure 1.**
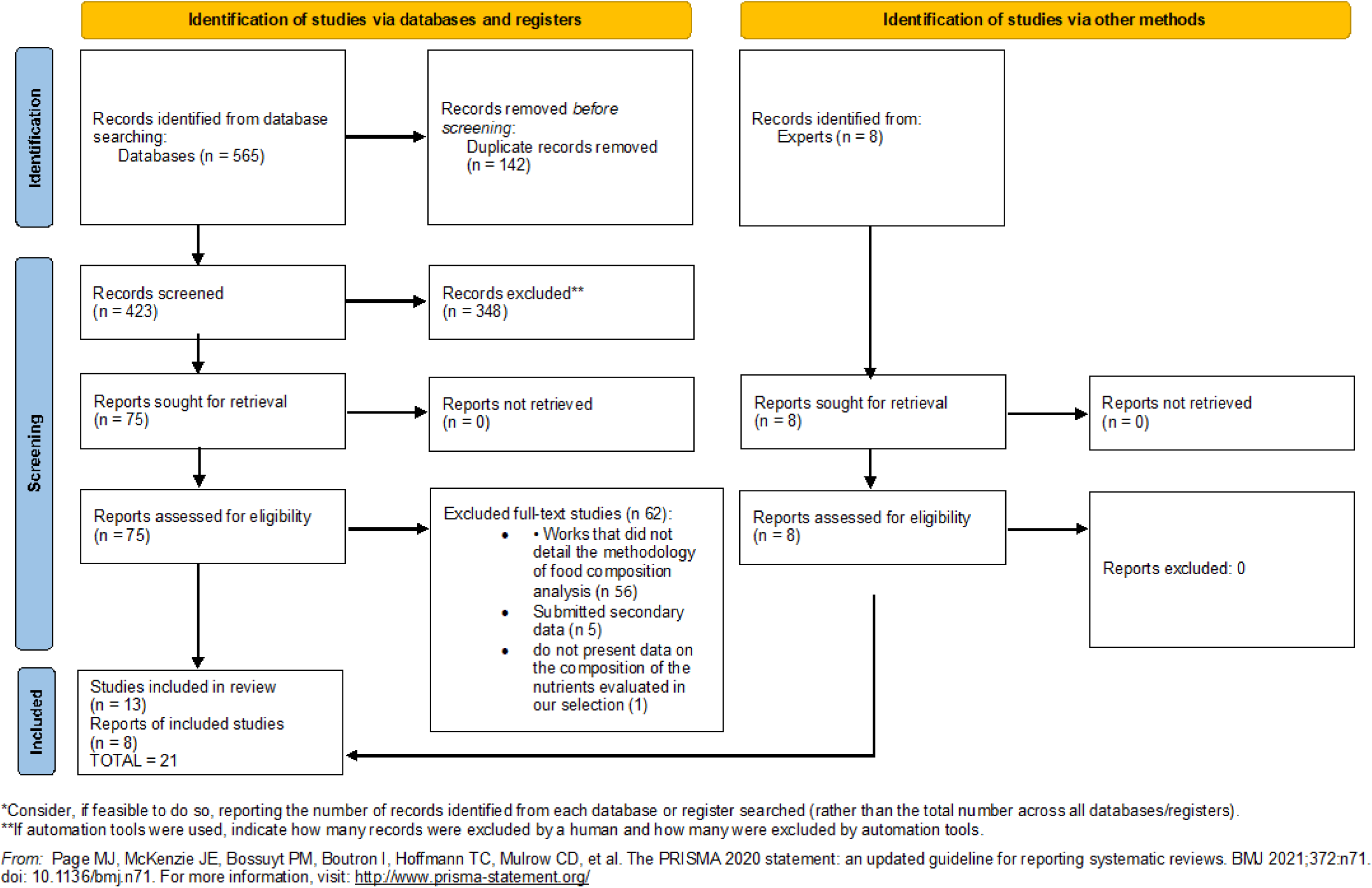
Flowchart of the study selection process.

### 3.2. Characteristics of the studies

The systematic review analyzed 26 distinct animal species, including 13 mammalian species, six bird species, and seven reptile species (see Supplementary Material 4). The studies covered sample collections from 11 countries: Italy, Germany, South Africa, Poland, the United States of America, New Zealand, Spain, Canada, Latvia, Peru, and Brazil. Italy, Spain, Poland, and South Africa were prominently featured for their recurring analyses of wildmeat. For a comprehensive understanding of the studies included in this review, Table 2 presents their main characteristics.

**Table 2.** Characterization of studies on the nutritional composition of wild animals. Names of animals collected are presented exactly as provided in the papers. Scientific names were updated for the analysis; for updated names, see Supplementary Material 4.

| n | Publication | Study objective | Key findings | Country of collection | Wild animals | Samples | Analyses performed | Study quality(29) |
| --- | --- | --- | --- | --- | --- | --- | --- | --- |
| 1 | Aguiar JPL. Tabela de composição de alimentos da Amazônia. ACTA AMAZONICA 26. 1996;121–6. | Analyze samples of wild animal meat obtained from public markets in the Amazon. | Analyses of the samples' composition were conducted. The results were compiled into a centesimal composition table, and a summary of the findings is available for consultation in this article. | Brazil | Cabeçudo (*Peltocephalus dumerilianus*); catitu (*Pecari tajacu*); cutia (*Dasyprocta leporina*); Iaçá (*Podocnemis sextuberculat*); jacaretinga (*Caiman crocodilus*); paca (*Agouti paca*); tartaruga (*Podocnemis expansa*); tracajá | 31 (muscle) + 4 (viscera) | Protein: Method Kjeldahl - AOAC (1984); Lipid: Soxhlet Method | High (6.50)(30) |
|  |  |  |  |  | (*Podocnemis unifilis*);<br>veado (*Mazama americana*) |  |  |  |
| 2 | Amici A, Cifuni GF, Contò M, Esposito L, Failla S. Hunting area affects chemical and physical characteristics and fatty acid composition of wild boar (*Sus scrofa*) meat. Rendiconti Lincei. 1o de novembro de 2015;26(3):527–34. | To examine how different geographic hunting areas influence the chemical composition and quality indices of wild boar meat. | The characteristics of wild animal meat varied due to environmental factors, including diet. Dietary diversity appears to influence aspects of the meat, such as color parameters, cooking losses, dry matter content, protein levels, and fatty acid profiles. These differences were observed in relation to the hunting area. | Italy | Wild boar (*Sus scrofa*) | 48 (Longissimus thoracis - muscle) | Protein: Method Kjeldahl - AOAC (1995);<br>Lipids: Folch et al. (1957);<br>Lipid Fraction: Method Soxhlet - IUPAC (1992). | Medium (5.50)(31) |
| 3 | Milczarek A, Janocha A, Niedzialek G, Zowczak-Romanowicz M, Horoszewicz E, Piotrowski S. Health-Promoting Properties of the Wild-Harvested Meat of Roe Deer (*Capreolus capreolus* L.) and Red Deer (*Cervus elaphus* L.) [Internet]. 2021 [citado 5 de setembro de 2024]. Disponível em: | Compare the proximate nutrient composition of roe deer and wild deer meat. | Regarding the macronutrient profile, no differences were observed with respect to sex, except for the fat content in females, which was higher than in males regardless of the species. As for micronutrients, the levels found are in accordance with the recommended values. | Poland | Roe deer (*Capreolus capreolus*), Red deer (*Cervus elaphus*) | 60 (semimembranosus muscles) | Protein: Kjeldahl (990.03); Lipids: Method Soxhlet (991.36); Lipid fractions: GC-FID; Methodology; Minerals: (Cd, Pb, Fe) PN-EN 14084 | High (8.33) |
|  | https://www.mdpi.com/2076-2615/11/7/2108 |  |  |  |  |  |  |  |
| 4 | Dannenberger D, Nuernberg G, Nuernberg K, Hagemann E. The effects of gender, age and region on macro- and micronutrient contents and fatty acid profiles in the muscles of roe deer and wild boar in Mecklenburg-Western Pomerania (Germany). Meat Sci. 10 de maio de 2013;94(1):39–46. | Investigate the effects of gender, age, and region on the macronutrients, micronutrients, and fatty acid profiles of wild boar and deer muscles. | The protein content in deer remained constant regardless of region, age, or sex; however, wild boar meat varied across these factors. Fat in deer varied by region, age, and sex, while in wild boar, they remained stable. Selenium and iron were not influenced by region, age, or sex, except for higher iron levels in older females. | Germany | Roe deer (*Capreolus capreolus*), Wild boar (*Sus scrofa*) | 203 (longissimus muscle) | Proteins and lipids: Meat Analyzer FoodScan™ (FOSS Analytic, Hillerød, Denmark); Lipid fractions: Gas chromatography analysis; Minerals: ICP-MS (Se, Cu, Fe, and Zn) | High (6.50) |
|  |  |  | Zinc concentrations varied according to region, age, and sex. |  |  |  |  |  |
| 5(32) | Fernandes HR, Deliza R, Neto OC, Silva CM, Albuquerque NI de, Martins TR, et al.<br>Effect of high hydrostatic pressure on the meat of collared peccaries (*Tayassu tajacu*) with different ages. Afr J Food Sci. 30 de setembro de 2022;16(9):215–25. | Evaluate the effects of high hydrostatic pressure on the physicochemical parameters of collared peccary meat. | High hydrostatic pressure tenderized the meat of adult collared peccaries and influenced meat quality. The results also showed changes in the protein profiles of animals of different ages subjected to different pressure levels. | Brazil | Peccary (*Pecari tajacu*) | 40 (muscle) | Proteins:<br>Method<br>Kjeldahl - AOAC (1995);<br>Lipids:<br>Method<br>Soxhlet - AOAC (1995);<br>Lipid fraction:<br>Bligh and Dyer (1959) | High (6.50)(33) |
| 6 | Gálvez C. H, Arbaiza T, Carcelén C. F, Lucas A. O. Revista de Investigaciones Veterinarias del Perú. 1999 [citado 5 de setembro de 2024].<br>VALOR NUTRITIVO DE LAS CARNES DE SAJINO (*Tayassu tajacu*), VENADO COLORADO (*Mazama americana*), MAJAZ (*Agouti paca*) Y MOTELO (*Georchelone denticulata*). | Determine the nutritional characteristics of wild animal meat. | Wild meats exhibited high protein content and low fat levels. | Peru | Sajino (*Tayassu pecari*), Motelo (*Chelonoidis denticulata*), Majaz (*Cuniculus paca*), Venado colorado (*Mazama americana*) | 20 (muscle) | Macronutrients and micronutrients<br>Proteins : Method Kjeldahl - AOAC, 1990;<br>Lipids: Method Soxhlet - .AOAC, 1990 | Medium (5.50) |
|  | Disponível em:<br>https://revistasinvestigacion.unmsm.edu.pe/index.php/veterinaria/article/view/6707 |  |  |  |  |  |  |  |
| 7(34)<br>) | Jarzyńska G, Falandysz J. Selenium and 17 other largely essential and toxic metals in muscle and organ meats of Red Deer (*Cervus elaphus*) — Consequences to human health. Environ Int. 1o de julho de 2011;37(5):882–8. | To determine the content and composition of micronutrients in red deer, relating them to meat intake rates, human nutritional needs, and associated health impacts. | The meats from muscles and organs of red deer exhibited high concentrations of copper, chromium, cobalt, manganese, selenium, and zinc. Additionally, they contained toxic trace elements such as mercury, lead, cadmium, and titanium. | Poland | Red Deer (*Cervus elaphus*) | 60 (20 muscle; 20 liver 20 de kidney) | Minerals: ICP-AES (Se) , ICP-MS (Ag, Ba, Cd, Co, Cr, Cs, Cu, Ga, Mn, Mo, Pb, Rb, Sb, Sr, Tl, V e Zn) | Medium<br>(4.50)(35) |
|  |  |  | The cadmium levels found in the meats exceed tolerance limits. |  |  |  |  |  |
| 8 | Johnson HE, Bleich VC, Krausman PR. Mineral deficiencies in tule elk, Owens Valley, California. J Wildl Dis. janeiro de 2007;43(1):61–74. | Assess the mineral content of horn, liver and forage to measure deficiencies and toxicities. | The levels of minerals found in the liver and horn varied between animals from two different locations. It was found that antler breakage in elk at one of the locations was related to a copper and phosphorus deficiency.<br>The levels of micronutrients found in forage varied according to location and season. | United States of America | Tule elk (*Cervus elaphus*) | 240 (liver) | Minerals:<br>Flame atomic absorption spectrophotometry (Cu e Mo); ICP-MS (Ca, Fe, Mg, Mn, S, P, Zn, Cu e Mo) | Medium (5.50)(36) |
| 9 | Lima AT.<br>Caracterização Fisico Quimica da Tartaruga da Amazônia de água proveniente de cativeiro e de habitat natural do estado do Amazonas.<br>Universidade Federal do Amazonas; 2009. | To evaluate the yield and chemical composition of *Podocnemis expansa* raised in commercial captivity and from natural habitats. | The meat evaluated in both systems showed a low fat content. And the free-living ones had a higher protein content. | Brazil | Tartaruga-da-amazônia (*Podocnemis expansa*) | 45 (muscle) + 15 (viscera) | Proteins : Method Kjeldahl ; Lipid (Bligh e Dyer,1959) | High (8.50)(37) |
| 10 | Lorenzo JM, Maggiolino A, Gallego L, Pateiro M, Serrano MP, Domínguez R, et al.<br>Effect of age on nutritional properties | Investigating the effects of slaughter age on the levels of macro and microminerals in wild deer meat. | Total fat and fraction contents were affected by the slaughter age, showing higher values in older animals. Regarding cholesterol, its levels decreased with | Spain | Red deer (*Cervus elaphus*) | 150 (muscle) | Lipid: Bligh e Dyer; Fraction lipid analysis: Gas chromatography; Minerals: ICP-OES, Ca, | High (8.50) |
|  | of Iberian wild red deer meat - Journal of the Science of Food and Agriculture [Internet]. 2019 [citado 5 de setembro de 2024]. Disponível em: https://scijournals.onlinelibrary.wiley.com/doi/abs/10.1002/jsfa.9334 |  | increasing slaughter age. |  |  |  | K, Mg, Na, P, Fe, Mn, Zn e Cu. |  |
| 11 | Cawthorn DM, Hoffman LC. The bushmeat and food security nexus: A global account of the | Determine the proximate and fatty acid composition of zebra meat. | The muscles had high protein content and low fat content. In the fat group, high concentrations of | South Africa | Zebra (*Equus quagga*) | 20 (longissimus lumborum muscle, longissimus thoracis et | Protein: Method Dumas - AOAC 992.15; Lipid: LEE; | High (7.33)(38) |
|  | contributions, conundrums and ethical collisions. Food Res Int. 1o de outubro de 2015;76:906–25. |  | saturated fatty acids were found. |  |  | lumborum) | Fraction lipid analysis: Gas chromatography; |  |
| 12 | Rudman M, Leslie AJ, van der Rijst M, Hoffman LC. Quality characteristics of Warthog (*Phacochoerus africanus*) meat. Meat Sci. 1o de novembro de 2018;145:266–72. | Evaluate the sensorial, physical and chemical attributes of wild boar meat. | Analysis of the sensory profile demonstrated that this meat has an undesirable aroma and flavor, described as sour, which was found in the meat of wild boars of all ages. Regarding macronutrient recommendations, these meats were considered healthy sources of protein and polyunsaturated fatty acids. | South Africa | Warthogs (*Phacochoerus africanus*) | 31 (Longissimus lumborum) | Protein: Method Dumas - 992.15; Lipids: LEE; Ashes (AOAC, 1992; AOAC, 2002a, 2002b) | High (8.0) |
| 13(39) | Neto JV, Bressan MC, Faria PB, Vieira JO e, Santana MTA, Kloster M. Composição centesimal e colesterol da carne de jacaré-do-pantanal (Caiman yacare Daudin 1802) oriundo de zoológico e habitat natural. Ciênc E | Determine the proximate composition and cholesterol of meat from the tail and back of alligators, from zoo breeding facilities and natural habitats. | Animals bred in captivity had a lower amount of fat and higher protein levels when compared to animals in their natural habitat. | Brazil | Alligator-swampland (*Caiman yacare*) | 12 (muscle) | Proteins : Method Kjeldahl - AOAC,1990; Lipids: Method Soxhlet - AOAC, 1990. | High (8.50) |
|  | Agrotecnologia.<br>agosto de<br>2006;30:701–6. |  |  |  |  |  |  |  |
| 14 | Landi N, Ragucci S,<br>Di Giuseppe AM,<br>Russo R, Poerio E,<br>Severino V, et al.<br>Nutritional profiling of<br>Eurasian woodcock<br>meat: chemical<br>composition and<br>myoglobin<br>characterization. J Sci<br>Food Agric.<br>2018;98(13):5120–8. | Evaluate the<br>nutritional aspects<br>of Eurasian chicken<br>meat. | Urian chicken meat<br>contains high<br>concentrations of high-<br>quality proteins, due to<br>its amino acid<br>composition and low fat<br>content. | Italy | Eurasian woodcock<br>(*Scolopax rusticola*) | 10 (entire chest<br>and leg muscle,<br>without skin) | Proteins :<br>Method<br>Kjeldahl;<br>lipids: Method<br>Soxhlet -<br>AOAC 991.36;<br>Lipid<br>fractions: GC-FID | High<br>(7.00)(41) |
|  | (40) |  |  |  |  |  |  |  |
| 15 | Pérez-Peña PE, Riveros-Montalván MS, Vargas-Arana G, Soria FD, Chumbe JV, Baca YB. Consumo, microbiología y bromatología de la carne silvestre durante la COVID-19 en Iquitos, Perú. Cienc Amaz Iquitos. | Know the microbiological and bromatological status of bush meat during the COVID-19 pandemic from two public markets in the Amazon. | Game meat had a low fat content and a high percentage of protein. The calcium content found in wild meat was higher than in domesticated meat. Phosphorus content was also significant in wild meat. | Peru | *Pecari tajacu*, *Tayassu pecari*, *Cuniculus paca*, *Chelonoidis denticulatus* | 16 (muscle) | Protein:Kjeldahl; Fat: Soxhlet; Vitamins: Chromatography; high pressure liquid (HPLC); Minerals: (zinc, sodium, potassium and | High (6.50)(26) |
|  | 2021;9(2):51–68. |  |  |  |  |  | iron) by<br>Absorption<br>Spectroscopy<br>Atomic (EAA)<br>and<br>Phosphorus by<br>UV Visible<br>Spectroscopy |  |
| 16 | Serrano MP,<br>Maggiolino A,<br>Landete-Castillejos T,<br>Pateiro M, Barbería<br>JP, Fierro Y, et al.<br>Quality of main types<br>of hunted red deer<br>meat obtained in Spain<br>compared to farmed | Assess the<br>combined impact of<br>country of origin<br>and type of<br>slaughter and season<br>on the quality and<br>nutritional value of<br>venison. | Wild deer from Spain<br>had a higher protein<br>concentration than meat<br>from animals raised in<br>New Zealand. The<br>calcium, sodium and<br>phosphorus levels found<br>in meat do not differ<br>between countries. New | Spain and<br>New<br>Zealand | Red deer (*Cervus*<br>*elaphus*) | 24 (Longissimus<br>thoracis et<br>lumborum) | Lipids: LEE;<br>Protein:<br>DUMAS | High (9.0) |
|  | venison from New Zealand. Sci Rep. 22 de julho de 2020;10(1):12157. |  | Zealand farm deer had higher concentrations of magnesium, iron, manganese, copper and lower concentrations of zinc when compared to wild deer from Spain. The nutrient profile of meats also varied depending on the season. Meat from winter hunts had higher levels of calcium, sodium, iron and zinc and lower levels of magnesium and phosphorus compared to<br>meat from summer hunts. |  |  |  |  |  |
| 17 | Sevillano-Caño J, Cámara-Martos F, Aguilar-Luque EM, Cejudo-Gómez M, Moreno-Ortega A, Sevillano-Morales JS. Trace Element Concentrations in Migratory Game Bird Meat: Contribution to Reference Intakes Through a | Determine the nutritional and toxicological content of game bird meat. | Iron and chromium contents were similar in all three bird species studied. Meat from thrush had the highest concentrations of copper. Pigeons and doves had the highest concentrations of zinc. The study concluded that the meat of migratory game birds is | Spain | Woodpigeon (*Columba palumbus*), Common turtledove (*Streptopelia turtur*), Thrush (*Turdus philomelos*) | 89 (breast; thigh) | Cu, Fe, Zn: flame atomic absorption spectroscopy (FAAS, Varian SpectraAA – model 50B); Cr, Co: Electrothermal Atomic Absorption Spectrometry | High (6.50) |
|  | Probabilistic Assessment. Biol Trace Elem Res. 1o de outubro de 2020;197(2):651–9. |  | an excellent dietary source of copper and iron, much better than other types of meat, such as non-wild birds and even red meat. |  |  |  | on an Agilent Model 240Z AA |  |
| 18 | Spiegelaar N, Martin ID, Tsuji LJS. Indigenous Subarctic Food Systems in Transition: Amino Acid Composition (Including Tryptophan) in Wild-Harvested and Processed Meats. Int J | Determine the amino acid and protein composition of wild meats and processed meats | The wild meats analyzed showed high concentrations of proteins and amino acids. | Canada | Moose (*Alces alces*), Canada Goose (*Branta canadensis*), Mallard Duck (*Anas platyrhynchos*) | 25 (muscle) | Proteins : Method Kjeldahl - AOAC 981.10; Amino acids: standardized hydrolysis of NaOH and HCl, and ultra-performance liquid | High (7.50) |
|  | Food Sci. 27 de junho<br>de<br>2019;2019:e7096416.<br>(42) |  |  |  |  |  | chromatograph<br>y |  |
| 19(43) | Strazdiņa V,<br>Jemeljanovs A, Šterna V. Nutrition Value of Wild Animal Meat. Proc Latv Acad Sci Sect B Nat Exact Appl Sci. 31 de outubro de 2013;67(4-5):373-7. | Compare the nutritional value of the meat of different types of wild animals with analogous domesticated animals. | Wild meat samples had a higher content of proteins, essential fatty acids, iron and zinc compared to non-wild animals. The highest cholesterol levels were found in wild boar (*Sus scrofa*). | Latvia | Elk (*Alces alces*), Deer (*Cervus elaphus*), Wild boar (*Sus scrofa*), Roe deer (*Capreolus capreolus*) | 54 (muscle) | Proteins :<br>Method<br>Kjeldahl;<br>Lipids:<br>Method<br>Soxhlet -<br>AOAC 991.36;<br>Lipid<br>fractions: Gas-<br>liquid chromatograph<br>y; Cholesterol:<br>Colorimetric<br>Blur;<br>Minerals: ICP-AES/ISO<br>6869-2002 | Medium<br>(5.00) |
| 20 | Zimmerman TJ, Jenks<br>JA, Leslie DM, Neiger<br>RD. Hepatic minerals<br>of white-tailed and<br>mule deer in the<br>southern Black Hills,<br>South Dakota. J Wildl<br>Dis. abril de<br>2008;44(2):341–50. | Assess hepatic<br>mineral<br>concentrations of<br>deer from different<br>habitats. | The liver mineral levels<br>found differed. The<br>differences were<br>associated with diet,<br>location and<br>reproductive stage. | United<br>States of<br>America | White-tailed deer<br>(*Odocoileus*<br>*virginianus*), Mule deer<br>(*Odocoileus hemionus*) | 83 (liver) | Minerais: ICP-AES Al, Sb,<br>As, Ba, B, Cd,<br>Ca, Cr, Co,<br>Cu, Fe, Pb,<br>Mg, Mn, Mo,<br>P, K, Se, Na,<br>S, Tl, Zn | Medium<br>(5.50) |
| 21(44) | Webb EC, Ryssen JBJ van, Erasmus MEA, McCrindle CME.<br>Copper, manganese, cobalt and selenium concentrations in liver samples from African buffalo (*Syncerus caffer*) in the Kruger National Park. J Environ Monit. 29 de novembro de 2001;3(6):583–5. | Evaluate the levels of micronutrients found in buffalo liver and in different regions. | The results demonstrated that there are significant differences in the amounts of micronutrients available in meat depending on the territory in which the buffalo is located. | South Africa | Buffalo (*Syncerus caffer*) | 311 (liver) | Minerals: ICP-AES copper, manganese and cobalt | High (6.67) |

### 3.3 Quality Analysis

During the quality analysis process, the agreement among the assessors was excellent (Fleiss’ Kappa = 1.00). No articles were rated as low quality, and 72% received a score indicating high quality (> 6.0 points) (see Supplementary Material 5). The highest ratings were given for procedures used in sample processing, description of analysis methods, and presentation of results with measures such as standard deviation or standard error. On the other hand, the criteria where the articles scored lower pertain to the exclusion of the scientific name of the species, as well as the absence of information on conflicts of interest and sources of funding for the studies.

### 3.4 General Comments on the Dataset

In the topic of data imputation, the techniques that showed the lowest SMAPE values for each nutrient demonstrated less absolute error and greater consistency, and were therefore selected for imputing the data. SMAPE values ranged from approximately 11.06% (protein) to 83.67% (selenium). For more details, see Supplementary Material 6.

### 3.5 General Comments on the Nutritional Profile of the Meats

All methods used in food composition analyses among the different studies are recognized by the AOAC (Association of Official Analytical Chemists), which supports the comparison of the obtained results (see Supplementary Material 7). The Supplementary Material 8 summarizes the mean and standard deviation values of the nutritional composition of wild animals included in this systematic review. Results of the statistical analyses are available in Supplementary Material 9.

Table 3 presents the main results, highlighting the pairs of comparisons where statistical differences were significant. We present the main results of this section by highlighting the following findings: differences in the nutritional composition of muscles among classes of wild animals, differences in the nutritional composition of visceral meat from different classes of wild animals, and differences in nutritional composition between muscle and visceral meats, including a comparison of nutrient profiles between these anatomical parts of wild animals.

**Table 3.** Nutritional composition in muscle and viscera of wildmeat, comprising mammals, birds, and reptiles, including original and imputed data. Pairs of comparisons with statistically significant differences are highlighted in black.

| Nutrient | Mammal<br>Viscera x<br>Mammal<br>Muscle | Mammal<br>Viscera x Bird<br>Muscle | Mammal<br>Viscera x<br>Reptile<br>Viscera | Mammal<br>Viscera x<br>Reptile Muscle | Muscle<br>Mammal x<br>Muscle Bird | Mammal<br>Muscle x<br>Reptile<br>Viscera | Mammal<br>Muscle x<br>Reptile Muscle | Bird muscle x<br>Reptile<br>Viscera | Bird Muscle x<br>Reptile Muscle | Reptile<br>Viscera x<br>Reptile Muscle |
| --- | --- | --- | --- | --- | --- | --- | --- | --- | --- | --- |
| Protein,<br>g | 23.26 x 23.15<br>(>0.05) | 23.26 x 23.46<br>(>0.05) | 23.26 x 16.33<br>(<0.01) | 23.26 x 21.34<br>(<0.01) | 23.15 x 23.46<br>(>0.05) | 23.15 x 16.33<br>(<0.05) | 23.15 x 21.34<br>(>0.05) | 23.46 x 16.33<br>(<0.01) | 23.46 x 21.34<br>(<0.01) | 16.33 x 21.34<br>(>0.05) |
| Fat, g | 1.37 x 1.27<br>(>0.05) | 1.37 x 2.42<br>(<0.05) | 1.37 x 5.7<br>(<0.05) | 1.37 x 2.02<br>(>0.05) | 1.27 x 2.42<br>(<0.01) | 1.27 x 5.7<br>(<0.01) | 1.27 x 2.02<br>(>0.05) | 2.42 x 5.7<br>(>0.05) | 2.42 x 2.02<br>(<0.05) | 5.7 x 2.02<br>(<0.05) |
| w6, g | 0.24 x 1.37<br>(<0.01) | 1.37 x 0.83<br>(>0.05) | 1.37 x 1.41<br>(>0.05) | 1.37 x 0.5<br>(<0.01) | 0.24 x 0.83<br>(<0.01) | 0.24 x 1.41<br>(<0.01) | 0.24 x 0.5<br>(>0.05) | 0.83 x 1.41<br>(>0.05) | 0.83 x 0.5<br>(>0.05) | 1.41 x 0.5<br>(>0.05) |
| Fe, mg | 14.51 x 3.21<br>(<0.01) | 4.41 x 14.51<br>(<0.01) | 14.51 x 15.55<br>(>0.05) | 14.51 x 5.39<br>(<0.01) | 3.21 x 4.41<br>(>0.05) | 3.21 x 5.39<br>(<0.01) | 3.21 x 5.39<br>(<0.01) | 3.21 x 15.55<br>(<0.01) | 4.41 x 5.39<br>(>0.05) | 15.55 x 5.39<br>(<0.01) |
| K, g | 0.13 x 0.21<br>(<0.05) | 0.13 x 0.27<br>(<0.01) | 0.13 x 0.06<br>(>0.05) | 0.13 x 0.21<br>(>0.05) | 0.21 x 0.27<br>(<0.01) | 0.21 x 0.06<br>(<0.05) | 0.21 x 0.21<br>(>0.05) | 0.27 x 0.06<br>(<0.01) | 0.27 x 0.21<br>(<0.05) | 0.06 x 0.21<br>(>0.05) |
| Mn, mg | 0.39 x 0.08<br>($<0.01$) | 0.39 x 0.12<br>($<0.01$) | 0.39 x 0.38<br>($>0.05$) | 0.39 x 0.14<br>($>0.05$) | 0.08 x 0.12<br>($>0.05$) | 0.08 x 0.38<br>($<0.01$) | 0.08 x 0.14<br>($<0.01$) | 0.12 x 0.38<br>($>0.05$) | 0.12 x 0.14<br>($>0.05$) | 0.38 x 0.14<br>($>0.05$) |
| Na, g | 0.1 x 0.11<br>($<0.01$) | 0.11 x 0.1<br>($>0.05$) | 0.1 x 0.09<br>($>0.05$) | 0.1 x 0.1<br>($>0.05$) | 0.11 x 0.1<br>($>0.05$) | 0.11 x 0.09<br>($<0.05$) | 0.11 x 0.1<br>($>0.05$) | 0.1 x 0.09<br>($>0.05$) | 0.1 x 0.1<br>($>0.05$) | 0.09 x 0.1<br>($>0.05$) |
| Zn, mg | 2.43 x 1.24<br>($<0.01$) | 3.33 x 0.86<br>($<0.01$) | 3.33 x 2.43<br>($>0.05$) | 3.33 x 1.24<br>($<0.01$) | 1.76 x 0.86<br>($<0.01$) | 1.76 x 2.43<br>($>0.05$) | 1.76 x 1.24<br>($<0.01$) | 0.86 x 2.43<br>($<0.01$) | 0.86 x 1.24<br>($>0.05$) | 2.43 x 1.24<br>($<0.01$) |
| Se, mcg | 57.19 x 10.61<br>($<0.01$) | 57.19 x 3.45<br>($<0.01$) | 57.19 x 81.17<br>($>0.05$) | 3.33 x 1.24<br>($<0.05$) | 10.61 x 3.45<br>($>0.05$) | 10.61 x 81.17<br>($>0.05$) | 10.61 x 19.37<br>($>0.05$) | 3.45 x 81.17<br>($<0.01$) | 3.45 x 19.37<br>($<0.01$) | 81.17 x 19.37<br>($>0.05$) |

### 3.6 Nutritional comparison: Nutritional Variation in Muscles of Mammals, Birds, and Reptiles

Our results demonstrate significant variations in the nutritional profile of meat depending on the analyzed class (mammal, bird, or reptile).

For reptiles, which were all aquatic species, their muscles showed over 60% higher concentrations of iron (p < 0.01) and over 75% higher concentrations of manganese (p < 0.01) compared to mammals. Additionally, reptile muscles exhibited selenium concentrations that were over 400% higher than those in bird muscles (p < 0.01) and nearly 60% higher fat concentrations than those in mammals.

Bird muscles stood out for having the highest concentrations of potassium compared to mammals (p < 0.01) and reptiles (p < 0.05). They also displayed elevated fat concentrations, which were over 90% higher than those found in mammal muscles (p < 0.01) and 20% higher than those in reptile muscles (p < 0.01). Furthermore, birds showed the highest concentrations of omega-6, surpassing those in mammals by over 200% (p < 0.01). There was no statistical difference in iron content between bird and mammal muscles; however, bird muscles had iron concentrations over 30% higher than those in mammals.

Mammal muscles exhibited the highest concentrations of zinc, with values over 100% higher than those found in bird muscles (p < 0.01) and over 40% higher than those recorded in reptile muscles (p < 0.01). Mammals also had the lowest fat concentrations, nearly 50% lower than in birds and almost 40% lower than in reptiles.

### 3.7 Nutritional comparison: Nutritional Variation in Viscera of Mammals and Reptiles

When comparing the nutritional content of viscera meat, we found that mammalian viscera had the highest protein concentrations compared to reptiles (p < 0.01). Conversely, reptilian viscera stood out for their high fat content, with concentrations over 300% higher than those found in mammals (p < 0.01).

### 3.8 Comparing Anatomical Parts: Nutritional Variation of Macro and Micronutrients in Viscera and Muscle of Mammals and Reptiles

Regarding the animal parts, the most frequent variations in macronutrients were related to fat content. We found that reptilian viscera exhibited higher fat content (5.37 g), nearly double (p < 0.05), and lower protein content (16.33 g) compared to muscle (2.02 g and 21.34 g, respectively). In mammals, the difference was significant only for omega-6, with viscera (1.37 g) showing over 400% higher content than muscles (0.24 g) (p < 0.01).

Regarding minerals, viscera generally had higher concentrations compared to muscles. Reptilian viscera showed almost 300% more iron (p < 0.01) and nearly 200% more zinc (p < 0.01) than their muscles. In mammals, viscera revealed iron and manganese concentrations over 300% higher (p < 0.01), selenium levels significantly higher (p < 0.01), and zinc concentrations almost 90% higher (p < 0.01) than those found in muscles. However, potassium content showed a different pattern, with muscle concentrations over 60% higher than those found in viscera (p < 0.01).

## 4. DISCUSSION

This study analyzes the nutritional composition patterns found in wild animal meat. Based on these findings, several important conclusions can be drawn: (i) wild animal meats represent significant sources of nutrients, which are often scarce in the diets of populations facing food insecurity; (ii) the viscera of these animals are particularly rich in minerals compared to muscles, similar to what is observed in meats from domesticated animals; and (iii) the nutritional profile, especially in terms of micronutrients, varies among different classes of animals.

Considering the interclass variation in both viscera and muscles, it is important to note that, although variations were observed, it is not possible to determine from the available data whether these variations are attributable to intrinsic characteristics of the class or to individual or environmental factors influencing the composition.

### 4.1 Wildmeat as Source of Nutrients

We compare the nutritional content of wildmeats with the DRI (Dietary Reference Intakes) to provide a reference for understanding the contribution of these wildmeats as a significant source of nutrients for people at different life stages (Supplementary Material 10). Wildmeats can be considered excellent sources of various nutrients. Analyses of the edible portions of muscles and viscera from these animals demonstrate their ability to provide high levels of proteins, as well as micronutrients such as iron, zinc, and selenium (see Supplementary Material 10). These elements are often deficient in the diets of vulnerable groups, such as women and children, particularly in IPLC contexts (45).

Nutritional deficiencies have significant health implications. Protein deficiency can lead to growth delays and induce hormonal and immunological changes (46). Iron deficiency is correlated with the development of anemia, (7) while zinc deficiency can lead to immune system dysregulation, compromised cognitive functions, growth deficits, and dermatological disorders (47). Selenium deficiency can compromise the effectiveness of immune system cells (48).

Our findings reveal that wild bird meat, in portions equivalent to a breast or thigh piece (100 g), can supply more than half of the daily iron needs for children and women, as well as more than half of the protein needed for women and nearly double the amount needed for children. Meanwhile, mammal meats, in a portion like a medium steak (100 g), provide about half of the protein and 40% of the iron needed for both children and women. Additionally, organs such as the liver and heart are even richer, potentially exceeding 200% of children’s iron and selenium needs. For reptiles, a portion of two small pieces of muscle (100 g) can meet over 200% of children’s iron needs and exceed 400% of their selenium needs. For women, the amounts of iron and selenium provided meet approximately 200% and 150% of their requirements, respectively.

These results highlight the potential of wild animal meats as sources of nutrients that are often deficient in the diets of populations facing food insecurity in developing countries. They contribute to improving dietary quality, making their consumption an important nutritional strategy for populations dependent on these resources (49). For this reason, either restricting access to wildmeats due to stringent conservation policies or defaunation of wild animal populations may have adverse nutritional impacts on these vulnerable human populations.

### 4.2 Viscera of Wild Animals are Important Reservoirs of Nutrients

Our findings indicated that meats derived from viscera have a substantially different nutritional composition compared to those from muscles. Both mammalian and reptilian viscera stood out for their highconcentrations of nutrients, surpassing muscular meats, particularly in terms of mineral content.

The composition of meat can vary considerably according to the anatomical part and tissue type of the animal (17). While meats derived from viscera are known for their high fat and micronutrient content, muscles are notable for their higher protein concentrations and lower levels of fat and micronutrients.

Despite variations in nutrient patterns between animal classes (reptiles and mammals), we observed that reptilian viscera demonstrated a pattern similar to that mentioned by, (17) with higher fat content, lower protein concentrations, and overall mineral concentrations nearly three times higher than in muscles. However, mammalian viscera, while exhibiting higher fat concentrations, particularly omega-6, and high concentrations of micronutrients, had protein concentrations similar to those found in muscles.

Furthermore, we observed a distinctive distribution of potassium compared to other minerals present in the viscera. Potassium concentrations in visceral meats were lower than in muscles for both reptiles and mammals. This disparity may be related to the physiological characteristics of these tissues, (17) as muscles require higher concentrations of potassium for muscle contraction.

Although wild animal viscera are important nutrient sources, we do not recommend promoting the preferential consumption of specific anatomical parts over others. Utilizing food holistically, a common practice among indigenous peoples and traditional communities, contributes to both sustainability, (45) and nutritional diversification through a complementarity mechanism (50). Furthermore, while viscera may contain high concentrations of certain nutrients, their proportion, both available and consumed, tends to be lower compared to other edible tissues of the animal.

Moreover, it is important to note that consuming viscera, despite being a rich source of essential nutrients, may pose risks due to higher levels of heavy metals compared to the muscles of animals. These levels can be higher in wild animal viscera and, in some cases, exceed recommended limits set by health organizations (51). Regular consumption of wildmeat can lead to a significant increase in exposure to heavy metals, such as cadmium and lead, in families consuming these meats (51). Thus, one of the recommendations is to limit the consumption of viscera, such as liver, especially in children.

Some studies indicate that certain nutrients can mitigate the toxic effects of heavy metals through chelation. For example, a study conducted in the Brazilian Amazon, where the local population is exposed to a diet rich in selenium and heavy metals such as methylmercury, revealed that selenium plays a chelating role, (52) potentially mitigating the toxic effects of methylmercury. This finding underscores the complexity of nutritional and environmental interactions, highlighting the importance of considering socio-ecological contexts in nutritional recommendations. While high selenium intake can lead to toxicity, (53) the case of the Amazonian population illustrates how excessive consumption of certain nutrients, under certain conditions, can offer protective effects. This highlights the need for a nuanced approach in nutritional guidelines. Selenium was one of the nutrients found in higher concentrations in the organ meats of wild animals, with 100 g of reptile and mammal organ meats providing over 100% of daily recommendations for children and adults.

### 4.3 The Nutritional Composition of Wildmeat Varies Among Classes, Not Necessarily Due to Animal Physiology Factors

Our comparison of nutrient content in muscles among species revealed a notable predominance of potassium, fat, and omega-6 in birds compared to other animal classes. Wild birds assessed in this study showed an average fat content of less than 2.5%. Despite being higher than other classes, this is lower than expected for non-wild birds, which typically have fat concentrations around 5%, possibly due to variations in physical activity levels (54). Fat content can also vary considerably depending on the bird species; for example, ostriches generally have lower fat content compared to chickens (55).

However, the results obtained with the birds in this study are enlightening in demonstrating that variations in nutrients may not be directly related to the taxonomic class of the animal. We observed that the muscles of wild mammals do not have higher iron concentrations compared to those of wild birds, indicated by a lack of statistically significant difference, a finding that differs from the pattern observed in domesticated animals (10). Based on these findings, we propose two hypotheses to explain the high concentrations of iron and other micronutrients in the meat of wild birds: (i) the distinct anatomy of birds and their migratory lifestyle, and (ii) the slaughter method to which free-ranging birds are subjected.

Within bird species, physiological characteristics associated with flight capability influence the nutritional profile of their meat. For flight, birds primarily use the pectoral muscles, responsible for wing movement (56). The continuous use of these muscles at high frequencies requires a high energy cost, leading to physiological adaptations to sustain prolonged flights (56). During migration periods, wild birds exhibit a significant increase in pectoral muscle mass and metabolic rates to sustain flight, allowing them to go long periods without feeding or drinking, primarily relying on fat oxidation for energy (57). This can also affect the low fat concentrations in their meats.

When comparing nutrient content in the breast and thigh meat of migratory wild birds, breast muscle has nearly double the iron content found in thigh meat, (13) In contrast, domesticated birds like turkey have lower iron concentrations in breast meat compared to thigh meat. For example, 100 g of turkey breast meat contains approximately 0.73 mg of iron, whereas the thigh contains 1.57 mg per 100g, (10) demonstrating a different nutrient profile compared to migratory wild birds.

The high iron concentration in wild bird meat may also be attributed to the slaughter method used. Typically, wild birds are shot in the chest, a highly vascularized area near nutrient-rich organs such as the liver and heart (13). This process can cause the leakage of visceral contents, increasing nutrient content, including iron, in the muscle tissues. The evidence presented on bird behavior and slaughter methods suggests that variations in nutrient levels may not be directly related to the animal’s taxonomic class.

The anatomy and migratory behavior of wild birds permits them to have a more diverse diet, directly influencing the nutritional composition of their meat (13). We can assume that birds primarily consuming seeds and oilseeds, which are naturally rich in fats, omegas, and minerals, (10) tend to have meat that is richer in these nutrients. In contrast, carnivorous and omnivorous birds have a distinct nutritional profile. These characteristics differ significantly from domesticated birds, which are raised in confinement and tend to feed mainly on agricultural feeds designed for rapid growth (58). Over time, soils used for conventional food production can experience nutritional imbalances and low fertility, especially in terms of micronutrients such as iron, zinc, and manganese (59). Consequently, food produced in conventional systems tends to have lower concentrations of these micronutrients, reflected in lower levels in domesticated bird meats.

Regarding reptiles, a significant finding was the high concentration of selenium in their muscles and viscera, much higher than those observed in birds and mammals. These findings may be linked to the physiological characteristics of reptiles, which have the ability to absorb nutrients not only from their diets but also directly from their surrounding aquatic environment (60). Reptiles can take in trace elements through their skin and other permeable membranes while submerged in water. This dual absorption mechanism contributes to the significant presence of trace elements, such as selenium, in their meat (60).

The reptiles analyzed in this study are aquatic and were collected mainly in the Brazilian Amazon, known for significant variations in soil and water composition, influenced by different organic matter and soil biota, both geographically (e.g. white, black and transparent rivers) and between dry and rainy periods (61). This region is known for high selenium concentrations in the soil, which play a crucial role in its absorption by local foods and water bodies. For example, Amazonian soils are responsible for the abundant production of Brazil nuts (*Bertholletia excelsa*), a significant source of selenium. Studies show considerable variations in selenium content in these nuts across different Amazonian regions, suggesting significant environmental influence on their concentration. These findings have important implications for understanding the nutritional composition of reptile meat, as selenium may be influenced by the diet of these animals in the Amazon region (62).

Our results demonstrated that wild mammals exhibited the highest concentrations of zinc compared to other animal classes, along with elevated levels of iron and potassium. These findings are consistent with, (26) who investigated wild deer meat and found that potassium, zinc, and iron were the most abundant nutrients. However, the content of these nutrients can vary within species depending on the animal’s life cycle. For instance, male deer may experience cyclic physiological osteoporosis related to rapid antler growth, decreasing zinc content in the bloodstream and consequently in all other body tissues (26). The results of this study highlight significant variations in the nutritional composition of meat not only between different classes of animals but also among species within the same class.

Finally, it is important to note that ecosystems vary considerably between countries, and the wide variations in nutritional profiles are also associated with the specificities from which these species were collected. The study encompassed nutritional composition data from 26 animal species, originating from 11 different countries. For this reason, nutrient content in these meats is subject to various factors, as nutrient distribution in the region varies significantly according to soil and water type and collection area (59).

Therefore, in our findings, birds stood out for their high concentrations of potassium, lipids, and omega-6 fatty acids, while reptiles exhibited exceptionally high levels of selenium, likely influenced by selenium-rich environments such as the Amazon. Wild mammals, on the other hand, distinguished themselves with high concentrations of zinc, iron, and potassium. These findings suggest that while physiological and metabolic factors may play a role, the nutritional composition of meat is strongly shaped by environmental factors such as diet and habitat. Therefore, it is important to consider these nuances when evaluating the nutritional value of meat from different species, rather than relying solely on taxonomic classifications.

### 4.4 Novelty of the Research

To the best of our knowledge, this is the first study to compile and analyze data concerning a large number of distinct species spanning three different classes in a single analysis. Notably, we have also introduced the technique of data imputation, presumably for the first time in the field of nutritional analysis of wildmeat. While data imputation has been previously employed in areas such as metabolomics for nutritional analyses? (63) its application in this context is innovative. Additionally, during our investigation, we identified a lack of appropriate tools to assess the methodological quality of research related to the nutritional composition of wild animals. Specific ethical and legal considerations in this field exacerbate this need. To address this limitation, we developed our own quality questionnaire (see Supplementary Material 2), which assisted us in evaluating the quality of the studies included in the review, considering the complexity of the resource in question.

Several limitations were encountered during the development of this study. One major challenge was the lack of access to original data from the reviewed articles, which may have compromised our ability to capture data variability. Additionally, the lack of specific information on covariates impacting composition in the analyzed studies limited our ability to adjust for these elements during tests and conduct subgroup analyses, preventing us from conducting a traditional meta-analysis. Furthermore, the small sample size of reptile meats demands caution in interpreting results related to this group.

We also observed high SMAPE (Symmetric Mean Absolute Percentage Error) values for certain nutrients, indicating reduced quality in the predictions considering the available data. This suggests the need for caution in interpreting these results. Despite this, we proceeded with the imputation process based on three main grounds: (1) the potential for significant natural variability, especially concerning wildmeats; (2) the scarcity of available data for this specific type of resource, which inherently may contribute to greater uncertainty in the forecasts made; and (3) relevant ethical considerations, including the difficulties associated with obtaining wild meat samples.

Despite these limitations, our research provides a detailed analysis of nutritional patterns in wild animals, which can underpin and guide future investigations in this field. The data obtained could assist in formulating more precise hypotheses, such as the impact of different slaughter methods on iron content in birds or the influence of the environment on selenium content in reptiles. Furthermore, we emphasize the importance for researchers, especially those focused on wildlife management, to enrich their databases with detailed information on variables influencing nutritional composition. This would allow for more comprehensive analyses, enhancing wildlife conservation efforts and maximizing the benefits derived from these resources, especially for IPLC.

## 5. CONCLUSION

Wildmeat play a crucial role as nutrient sources for diverse human populations. Findings from this study reveal significant nutrient variations among different animal classes and anatomical parts, underscoring the ability of game meat to provide essential nutrients, especially micronutrients often deficient in the diets of vulnerable groups such as children and women of childbearing age who are particularly susceptible to the effects of malnutrition. Moreover, this study introduces the use of data imputation methodology as an alternative to navigating scarcity of data on wild foods. However, it is important to note that current primary data limitations hinder the precise determination of whether these differences are due to inherent characteristics of animal classes or to individual and environmental factors affecting nutritional composition. Future research that directly analyzes the nutritional composition of wildmeat is needed to clarify the causes of these variations.

## Data Availability

All data is available in Supplementary Material 3. Scripts for the analysis are available at https://github.com/eliasjacob/paper_nutritional_composition_wildmeat

---

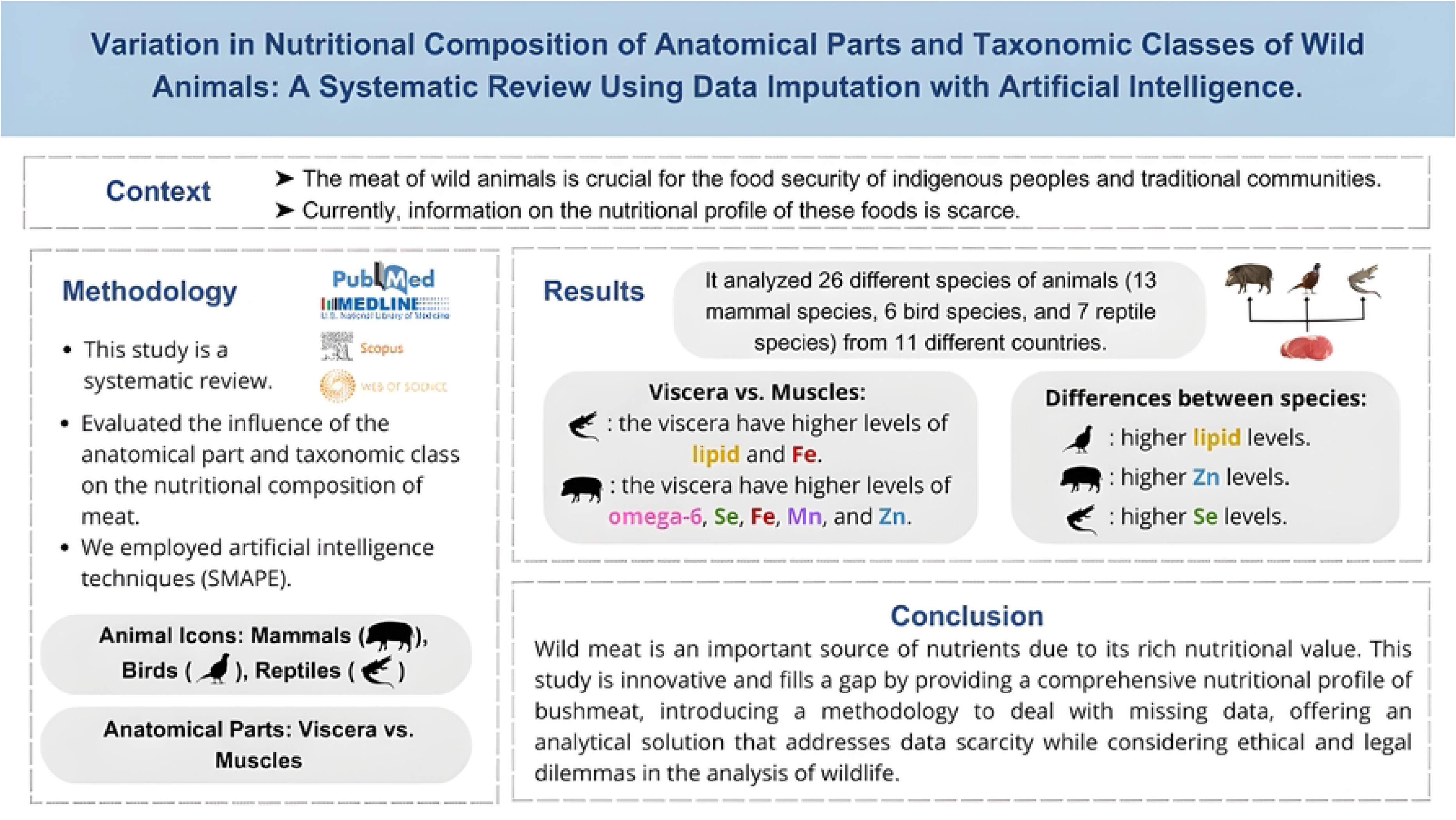

